# Comparative Efficacy and Safety of Left Atrial Appendage Occlusion Versus Direct Oral Anticoagulants in Atrial Fibrillation: A Systematic Review and Meta-Analysis

**DOI:** 10.1101/2025.09.21.25336272

**Authors:** Manish Juneja, Deep Patel, Manasi Garde, Aadhiti Kandula, Maria Elizabeth John, Parag N Patel, Goranti Shreya Reddy, Vivek Shekar, Rakhshanda Khan, Harshawardhan Dhanraj Ramteke, Syeda Hafsa Noor-Ain

## Abstract

**Introduction:** Left atrial appendage occlusion (LAAO) has emerged as an alternative to direct oral anticoagulants (DOACs) for stroke prevention in atrial fibrillation (AF), particularly in patients with contraindications to long-term anticoagulation. However, comparative efficacy and safety remain uncertain.

**Methods:** We conducted a systematic review and meta-analysis of studies published up to September 2025. A total of 395 studies were screened, and 14 met eligibility criteria, encompassing 683,659 patients. Data on baseline demographics, thromboembolic outcomes, bleeding complications, and mortality were extracted. Pooled estimates were calculated using random-effects models in Stata 18.0. Risk of bias was assessed with ROBINS-I and RoB 2.0, and certainty of evidence was graded using GRADE.

**Results:** Of the total population, 14,931 patients underwent LAAO and 640,888 received DOACs. The cohort included 287,031 males and 339,818 females, with hypertension present in 8262 LAAO and 189,627 DOAC patients, CKD in 4256 vs 122,521, and diabetes in 7422 vs 284,861, respectively. LAAO patients had a mean CHA DS -VASc score of 4.58 compared to 4.92 in DOAC patients. Meta-analysis showed no significant differences between LAAO and DOACs in all-cause mortality (RR 0.08; 95% CI –0.77 to 0.92), cardiovascular mortality (RR 0.67; 95% CI –0.60 to 1.94), thromboembolic events (RR 0.47; 95% CI –0.83 to 1.77), or major bleeding (RR 0.58; 95% CI –0.71 to 1.87).

**Conclusion:** LAAO provides stroke prevention comparable to DOACs without significant differences in mortality or bleeding. LAAO may represent an appropriate option in AF patients with high bleeding risk or contraindications to anticoagulation. Further randomized trials are warranted to refine patient selection and confirm long-term outcomes.

## Introduction

Atrial fibrillation (AF) is the most common sustained arrhythmia encountered in clinical practice, affecting millions worldwide and contributing substantially to the global burden of cardiovascular disease [1]. Its prevalence rises steeply with age, and with the growing elderly population, the incidence of AF is expected to increase further in the coming decades [2]. One of the most serious complications of AF is thromboembolic stroke, with the left atrial appendage recognized as the predominant site of thrombus formation in non-valvular AF [3]. Effective stroke prevention therefore lies at the heart of AF management strategies.

For decades, oral anticoagulation has remained the standard approach to reduce stroke risk in AF [4]. The introduction of direct oral anticoagulants (DOACs) offered important advantages over vitamin K antagonists, including more predictable pharmacokinetics, fewer food and drug interactions, and the elimination of routine monitoring [5]. Randomized controlled trials have firmly established their efficacy and safety across a wide spectrum of AF populations, making DOACs the first-line therapy in most guidelines [6]. However, anticoagulation is not without drawbacks. A proportion of patients remain at unacceptably high risk of bleeding, others have contraindications to long-term anticoagulation, and adherence challenges persist in real-world practice [7]. These limitations have stimulated the search for non-pharmacologic strategies to mitigate stroke risk in AF.

Left atrial appendage occlusion (LAAO) has emerged as a promising alternative [8]. By mechanically sealing the left atrial appendage, LAAO aims to eliminate the nidus for thrombus formation and thereby reduce embolic risk without the need for indefinite systemic anticoagulation [9,10]. Several devices, including the Watchman and Amplatzer Amulet, have been developed and increasingly adopted in clinical practice. Initial randomized trials comparing LAAO with warfarin demonstrated non-inferiority for stroke prevention and a reduction in hemorrhagic events over long-term follow-up [11]. Yet, these trials pre-dated the widespread use of DOACs, leaving unanswered the question of how LAAO compares with current standard oral anticoagulant therapy.

In this context, observational studies have become particularly valuable [12]. Over the past decade, registry data and cohort analyses have begun to address this evidence gap by comparing outcomes between patients receiving LAAO and those treated with DOACs [13]. Several large multicenter registries have suggested that LAAO may provide stroke prevention comparable to DOACs while conferring lower rates of major bleeding and, in some reports, improved survival [14]. Propensity-matched analyses from national health databases have reinforced these observations, demonstrating consistent reductions in composite outcomes driven largely by fewer bleeding events and deaths. Smaller single-center studies, while limited in sample size, have offered additional insights into patient selection, procedural safety, and device-related complications.

Despite this growing body of evidence, uncertainties remain [15]. Observational studies are inherently subject to residual confounding and selection bias, as patients referred for LAAO are often those at highest bleeding risk or with prior hemorrhagic events, while DOAC cohorts may represent a broader or healthier population. Variability in patient characteristics, device types, operator experience, and follow-up duration further complicates interpretation. Outcome definitions are not uniform across studies, with differences in how ischemic stroke, systemic embolism, and major bleeding are adjudicated. Furthermore, while some analyses suggest survival benefits with LAAO, others have not reproduced these findings, highlighting the need for systematic synthesis.

The clinical stakes are high. If LAAO provides comparable stroke prevention with lower bleeding risk, it could be considered not only in patients unable to tolerate anticoagulation but also as an alternative in broader groups [16]. Conversely, if DOACs continue to provide superior net clinical benefit, their primacy would be reinforced. To date, no single study has provided a definitive answer, and randomized evidence directly comparing LAAO with DOACs remains scarce. Against this backdrop, a comprehensive meta-analysis of observational studies is warranted to consolidate existing data, quantify treatment effects, and provide clarity for clinicians and policymakers.

The aim of the present study is therefore to systematically evaluate and synthesize observational evidence comparing LAAO with DOACs in patients with atrial fibrillation. Specifically, this meta-analysis will assess the relative risks of ischemic stroke, major bleeding, all-cause mortality, and composite clinical endpoints. By integrating findings across diverse settings and populations, this work seeks to generate robust estimates of comparative effectiveness, explore heterogeneity across subgroups, and identify gaps in the evidence that should inform the design of future randomized trials. Ultimately, the goal is to provide an evidence-based framework to guide clinical decision-making in this evolving field of AF management.

## Methods

### Literature Search

A comprehensive search was performed across PubMed, Embase, Cochrane Library, and Web of Science from database inception to September 2025, without language restrictions. Search terms combined controlled vocabulary and free text for “left atrial appendage occlusion,” “LAAO,” “Watchman,” “Amulet,” “direct oral anticoagulants,” “DOAC,” “NOAC,” “dabigatran,” “rivaroxaban,” “apixaban,” “edoxaban,” and “atrial fibrillation,” linked with Boolean operators. Reference lists of eligible articles and relevant reviews were hand-searched to identify additional studies. Both published and ahead-of-print records were considered. Observational studies directly comparing LAAO with DOAC therapy in atrial fibrillation were included, assessing outcomes of ischemic stroke, major bleeding, and all-cause mortality. Prisma guidelines were followed and registered with Prospero with number CRD420251152538 [17]

### Study Selection and Data Extraction

All titles and abstracts retrieved from the search were independently screened by two reviewers to identify potentially relevant studies. Full texts of selected articles were then assessed for eligibility according to predefined inclusion and exclusion criteria. Observational studies (prospective or retrospective cohorts, registry analyses, or case–control studies) directly comparing left atrial appendage occlusion (LAAO) with direct oral anticoagulants (DOACs) in patients with atrial fibrillation were eligible. Studies without a direct comparator group, those including only warfarin or mixed anticoagulation strategies without subgroup data, editorials, reviews, and case series with fewer than 10 patients were excluded. Disagreements were resolved through consensus or third-reviewer adjudication.

Data extraction was performed independently by two reviewers using a standardized form. Extracted variables included study characteristics (author, year, country, study design, sample size, and follow-up duration), patient demographics (age, sex distribution, comorbidities, CHA DS -VASc and HAS-BLED scores), intervention details (device type for LAAO, specific DOAC agent and dosing), and clinical outcomes (ischemic stroke or systemic embolism, major bleeding, all-cause mortality, and composite endpoints). When available, hazard ratios, relative risks, odds ratios, and their 95% confidence intervals were collected. If effect measures were not reported, raw event data were extracted to allow calculation. For studies reporting multiple models, effect estimates adjusted for the greatest number of confounders were preferred. In cases of missing or unclear data, study authors were contacted for clarification. Where overlapping populations were identified, the study with the longest follow-up or most comprehensive reporting was included. All extracted data were cross-checked for accuracy before synthesis.

#### Statistical Analysis

All statistical analyses will be conducted in accordance with best practices for meta-analyses of observational studies. Effect estimates will be expressed as relative risks (RRs), or odds ratios (ORs) with corresponding 95% confidence intervals (CIs). When available, the most fully adjusted HRs will be extracted and pooled. If adjusted estimates are not provided, crude event data will be used to calculate risk ratios.

Given the anticipated clinical and methodological heterogeneity among included studies, a random-effects model (DerSimonian–Laird method) will be applied. Statistical heterogeneity will be quantified using the I² statistic, with thresholds of 25%, 50%, and 75% considered low, moderate, and high heterogeneity, respectively. Cochran’s Q test will also be performed, with a p-value <0.10 indicating potential heterogeneity.

Subgroup analyses will be planned based on study design (prospective vs retrospective), device type (Watchman vs Amulet), DOAC type (dabigatran, rivaroxaban, apixaban, edoxaban), baseline bleeding risk (e.g., HAS-BLED score categories), and follow-up duration. Sensitivity analyses will be conducted by excluding studies at high risk of bias and by applying fixed-effects models to test the robustness of the findings.

Publication bias will be assessed through visual inspection of funnel plots and quantitatively using Egger’s regression test and Begg’s test, when ≥10 studies are available. If funnel plot asymmetry is detected, the trim-and-fill method will be applied to estimate the influence of unpublished studies.

All analyses will be performed using Stata version 18.0 (StataCorp LLC, College Station, TX, USA). A two-sided p-value <0.05 will be considered statistically significant.

#### Risk of Bias Assessment

The methodological quality of included observational studies will be assessed using the **ROBINS-I (Risk Of Bias In Non-randomized Studies of Interventions) tool [18]**. This evaluates risk of bias across seven domains: confounding, selection of participants, classification of interventions, deviations from intended interventions, missing data, measurement of outcomes, and selection of the reported result. Each domain will be graded as “low,” “moderate,” “serious,” “critical,” or “no information.” Two reviewers will independently assess each study, with disagreements resolved by consensus or consultation with a third reviewer. A summary risk-of-bias table and traffic-light plots will be generated to provide a visual overview of study quality.

Sensitivity analyses will be conducted by excluding studies judged to be at high or critical risk of bias to determine their influence on pooled estimates.

#### Certainty of Evidence (GRADE)

The certainty of evidence for each main outcome (ischemic stroke, major bleeding, all-cause mortality, and composite endpoints) will be evaluated using the **GRADE (Grading of Recommendations Assessment, Development and Evaluation) approach [19]**. The GRADE framework considers risk of bias, inconsistency, indirectness, imprecision, and publication bias. Observational studies start as “low-certainty” evidence but can be upgraded based on factors such as large effect sizes, evidence of a dose-response gradient, or when all plausible confounding would reduce the observed effect. Certainty of evidence will be rated as **high, moderate, low, or very low**, and “Summary of Findings” tables will be developed to present effect estimates alongside the GRADE certainty ratings. This structured approach will ensure transparent interpretation of the strength and reliability of the evidence, providing guidance for clinical practice and future research.

## Results

### Demographics and Baseline Characteristics

A total of **395 studies** were screened, of which **14 observational studies** fulfilled the inclusion criteria and were included in the final meta-analysis [20–33]. These studies comprised **683,659 patients**, with **14,931 (2.2%)** undergoing left atrial appendage occlusion (LAAO) and the remainder treated with direct oral anticoagulants (DOACs). The overall population included **287,031 males (45.2%)** and **339,818 females (49.7%)**. Baseline risk profiles were high across both groups. Patients undergoing LAAO had a mean **HAS-BLED score of 3.39 ± 1.31** and a **CHA**D**DS**D**-VASc score of 4.58 ± 1.62**, which were comparable to those in the DOAC-treated atrial fibrillation (AF) population (**3.41 ± 1.42** and **4.92 ± 1.63**, respectively). Hypertension was present in **8,262 (55.3%)** of the LAAO group and **189,627 (29.6%)** of the DOAC group. Similarly, **chronic kidney disease (CKD)** was more frequent in LAAO patients (**4,256; 28.5%**) compared with the AF cohort (**122,521; 19.1%**). The prevalence of **diabetes mellitus** was **7,422 (49.7%)** in LAAO versus **284,861 (44.4%)** in AF, while **heart failure** occurred in **6,216 (41.6%)** and **258,708 (40.4%)** patients, respectively. Patients selected for LAAO also had higher rates of prior events. A history of **transient ischemic attack (TIA)** was reported in **3,134 (21.0%)** LAAO patients and **94,740 (14.8%)** in AF. Likewise, **previous bleeding** was more common in the LAAO cohort (**6,503; 43.5%**) compared with AF patients on DOAC therapy (**170,677; 26.6%**). Taken together, these findings confirm that patients undergoing LAAO generally represent a **higher-risk clinical profile**, with more comorbidities and prior adverse events, despite broadly similar risk scores compared with those treated with DOACs. Table S1 and Figure 1.

**Figure 1.**
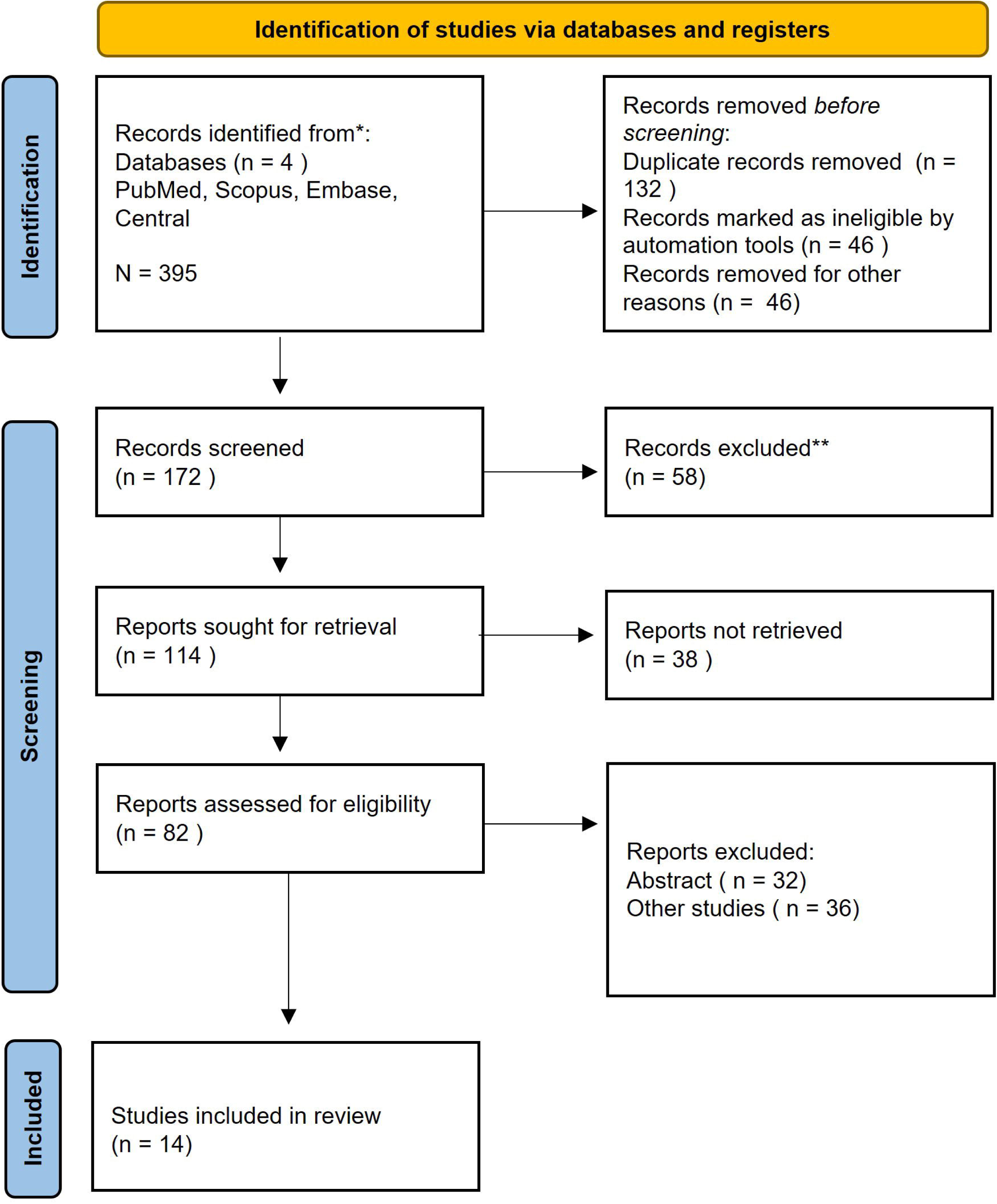
PRISMA Flow Diagram

**Figure 2.**
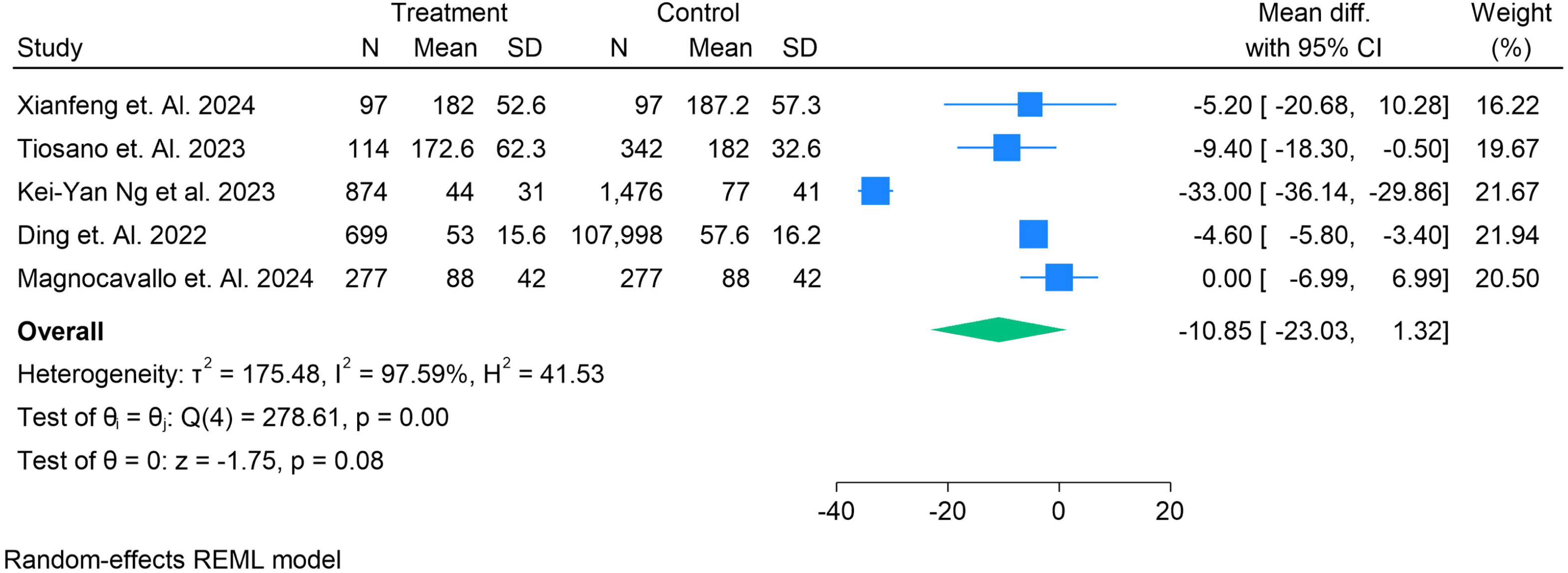
Mean Difference in the time taken by the procedure to complete in LAAO vs DOAC

### Procedural Characteristics

Five studies (n = 99,059 patients) reported on procedure completion time in LAAO compared with DOAC therapy. The pooled analysis using a random-effects REML model demonstrated **no statistically significant difference** between groups (mean difference: –10.85 minutes; 95% CI: –23.03 to 1.32; p = 0.08). Significant heterogeneity was observed (I² = 97.6%, τ² = 175.48, p < 0.001). Subgroup differences were evident across studies, with two trials (Tiosano et al. 2023, Kei-Yan Ng et al. 2023) reporting shorter procedural times for LAAO, while others (Xianfeng et al. 2024, Ding et al. 2022, Magnocavallo et al. 2024) showed no significant difference. These findings indicate that procedure time is largely comparable between LAAO and DOAC groups, with variability driven by study design, operator experience, and procedural complexity.

#### Bleeding Outcomes (BARC-defined Events)

Five studies reported on bleeding outcomes according to the Bleeding Academic Research Consortium (BARC) criteria in patients treated with LAAO compared with DOACs. The pooled analysis using a random-effects REML model demonstrated no statistically significant difference between groups (log odds ratio: 0.44; 95% CI: –0.36 to 1.25; p = 0.28).

There was substantial heterogeneity across studies (I² = 92.8%, τ² = 0.64, p < 0.001). Individual study results were variable: Ding et al. (2022) and Kei-Yan Ng et al. (2023) showed significantly higher BARC bleeding with LAAO, while Paiva et al. (2021) and Aarnik et al. (2024) reported no significant difference. Tiosano et al. (2023) demonstrated a modest increase in events among LAAO patients. Overall, these findings suggest that while LAAO may not consistently reduce BARC bleeding compared with DOACs, variation between studies highlights the influence of population selection, event definitions, and follow-up duration. Figure 3. Forest plot showing the number of people with BARC bleeding events in LAAO versus DOAC groups. The size of the squares represents study weight, and horizontal lines indicate 95% confidence intervals. The pooled effect was derived using a random-effects REML model.

**Figure 3.**
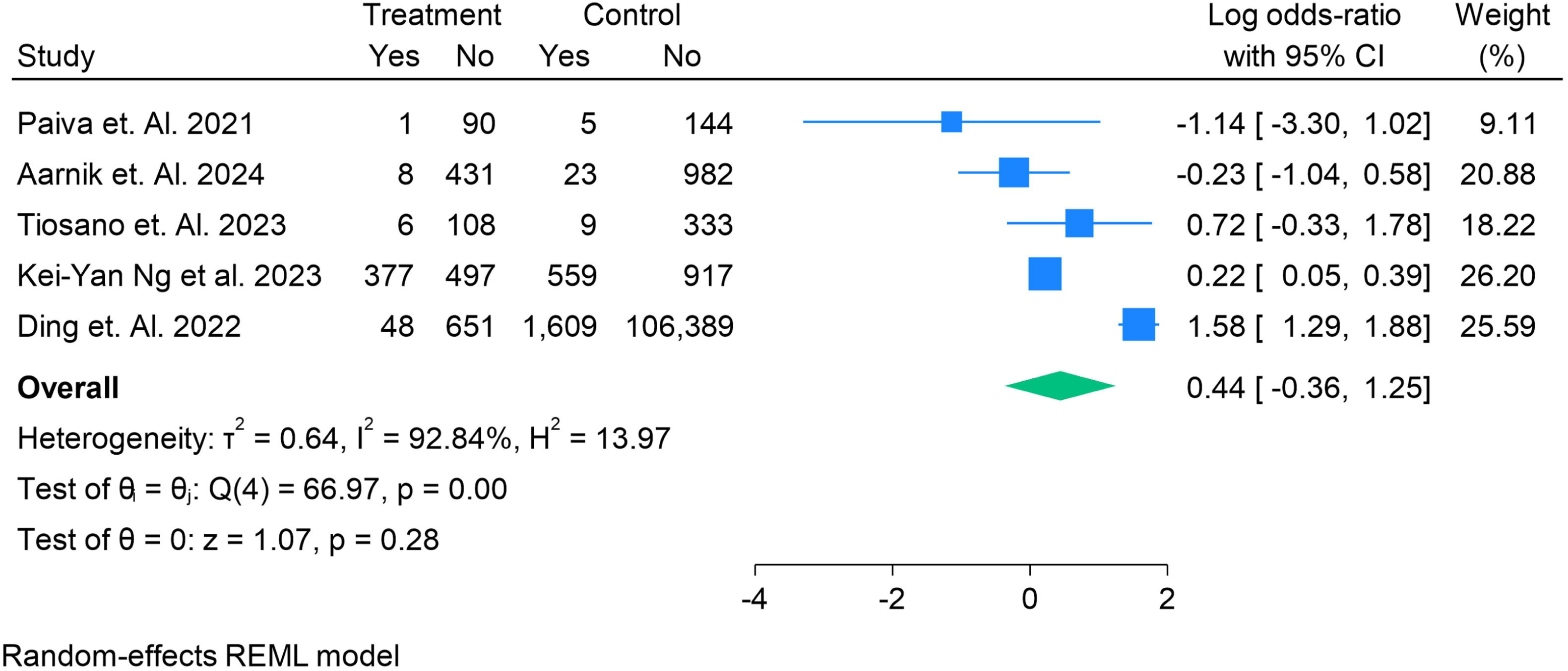
Number of People in BARC in the LAAO vs DOAC

#### Bleeding Outcomes (ISTH-defined Events)

Four studies reported bleeding outcomes according to the International Society on Thrombosis and Haemostasis (ISTH) criteria in patients treated with LAAO compared with DOACs. The pooled analysis using a random-effects REML model showed **no significant difference** between the two groups (log odds ratio: –0.15; 95% CI: –0.95 to 0.66; p = 0.72). Moderate heterogeneity was observed (I² = 50.4%, τ² = 0.32, p = 0.10). While Magnocavallo et al. (2024) reported a significantly lower rate of ISTH bleeding in the LAAO group, Paiva et al. (2021), Xianfeng et al. (2024), and Tiosano et al. (2023) demonstrated no significant differences. Overall, these findings suggest that ISTH-defined bleeding risk does not differ substantially between LAAO and DOAC therapy, although heterogeneity highlights differences in study populations and follow-up durations. **Figure 4**. Forest plot showing the number of patients with ISTH-defined bleeding events in LAAO versus DOAC groups. The size of the squares represents study weight, and horizontal lines indicate 95% confidence intervals. The pooled effect was estimated using a random-effects REML model.

**Figure 4.**
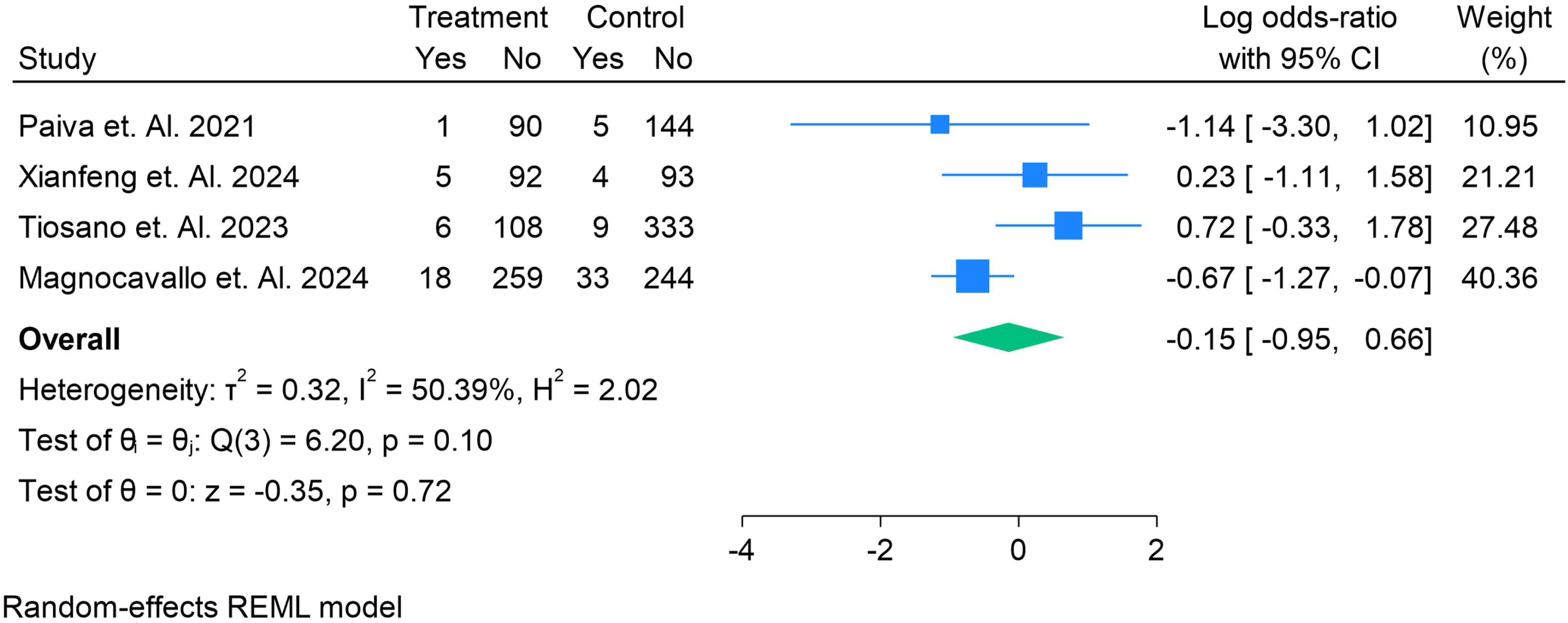
Number of People in ISTH in the LAAO vs DOAC

#### Periprocedural Bleeding

Five studies reported on the incidence of periprocedural bleeding in patients undergoing LAAO compared with those receiving DOAC therapy. The pooled analysis using a random-effects REML model demonstrated **no significant difference** between groups (log odds ratio: –0.42; 95% CI: –0.94 to 0.10; p = 0.12). Importantly, **no heterogeneity** was observed across included studies (I² = 0.0%, τ² = 0.00, p = 0.65), suggesting consistent findings. While Paiva et al. (2021), Xianfeng et al. (2024), and Magnocavallo et al. (2024) showed lower odds of periprocedural bleeding with LAAO, Osmancik et al. (2022) reported a non-significant increase, and Aarnik et al. (2024) demonstrated comparable results between groups. Overall, these findings indicate that LAAO does not significantly alter periprocedural bleeding risk compared with DOAC therapy, and the consistency across studies strengthens this conclusion. **Figure 5**. Forest plot showing the odds of periprocedural bleeding in patients undergoing LAAO compared with DOAC therapy. The size of the squares reflects the relative weight of each study, and horizontal lines indicate 95% confidence intervals. The pooled effect was estimated using a random-effects REML model.

**Figure 5.**
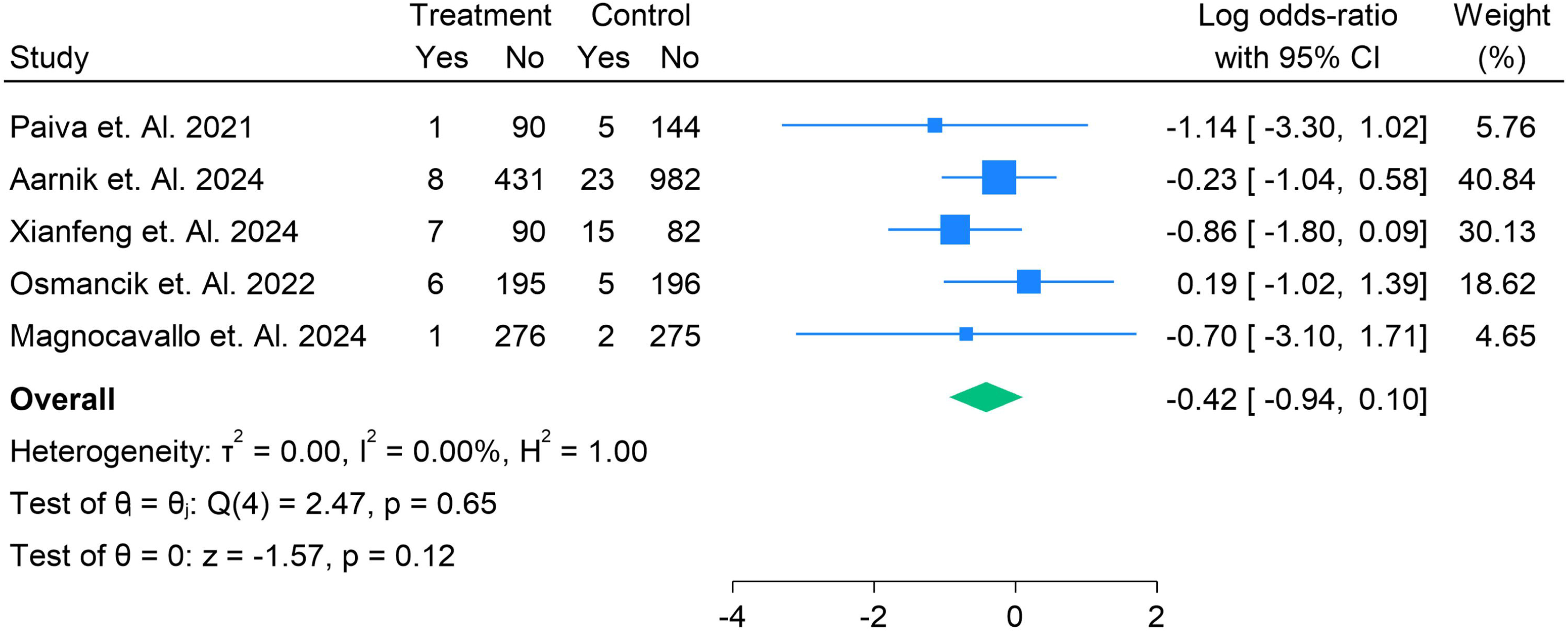
The Odds of the sample having the Peri-Procedural Bleeding in LAAO vs DOACs

#### Thrombus Events

Four studies reported the occurrence of thrombus events in patients undergoing LAAO compared with those receiving DOAC therapy. The pooled analysis using a random-effects REML model demonstrated **no significant difference** between the two groups (log odds ratio: 0.99; 95% CI: –2.12 to 4.11; p = 0.53). However, there was **substantial heterogeneity** (I² = 94.9%, τ² = 9.39, p < 0.001). Ding et al. (2022) showed a markedly higher odds of thrombus events in the LAAO group (log OR 5.18; 95% CI: 4.79–5.57), whereas Xianfeng et al. (2024) suggested a protective effect (log OR –2.16; 95% CI: –4.25 to –0.06). Paiva et al. (2021) and Magnocavallo et al. (2024) reported no significant differences between groups. Overall, these findings indicate **no consistent evidence of difference in thrombus events between LAAO and DOAC therapy**, although between-study variability suggests the need for standardized definitions and longer follow-up in future investigations. **Figure 6**. Forest plot showing the odds of thrombus events in patients treated with LAAO compared with DOAC therapy. The size of the squares corresponds to study weight, with horizontal lines representing 95% confidence intervals. Pooled estimates were generated using a random-effects REML model.

**Figure 6.**
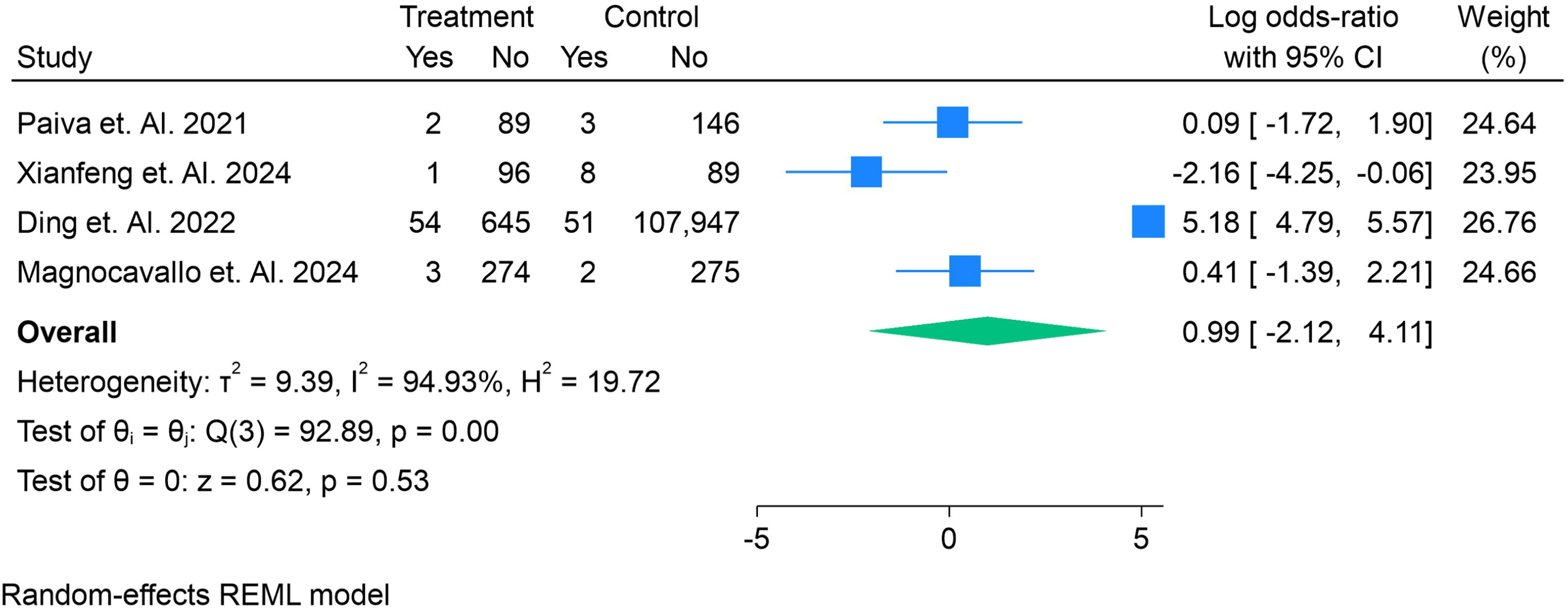
The Odds of the sample having the Thrombus Events in LAAO vs DOACs

#### Pericardial Effusion

Six studies evaluated the odds of pericardial effusion in patients undergoing LAAO compared with those treated with DOACs. The pooled analysis using a random-effects REML model showed **no significant difference** between groups (log odds ratio: 0.93; 95% CI: –0.69 to 2.55; p = 0.26). There was **considerable heterogeneity** across studies (I² = 89.8%, τ² = 3.54, p < 0.001). Turagam et al. (2017) reported a significantly higher incidence of pericardial effusion with LAAO (log OR 4.64; 95% CI: 3.32–5.96), while Xianfeng et al. (2024) and Kei-Yan Ng et al. (2023) demonstrated a nonsignificant trend towards increased risk. In contrast, Sigusch et al. (2025) and Kim et al. (2025) found no meaningful difference, and Magnocavallo et al. (2024) reported wide confidence intervals with uncertain effect. Overall, these findings suggest that **LAAO may not significantly increase the risk of pericardial effusion compared with DOACs**, but variability across studies underscores the influence of procedural experience, device type, and patient selection. **Figure 7**. Forest plot showing the odds of pericardial effusion in patients treated with LAAO compared with DOAC therapy. Square sizes correspond to study weight, and horizontal lines represent 95% confidence intervals. The pooled estimate was obtained using a random-effects REML model.

**Figure 7.**
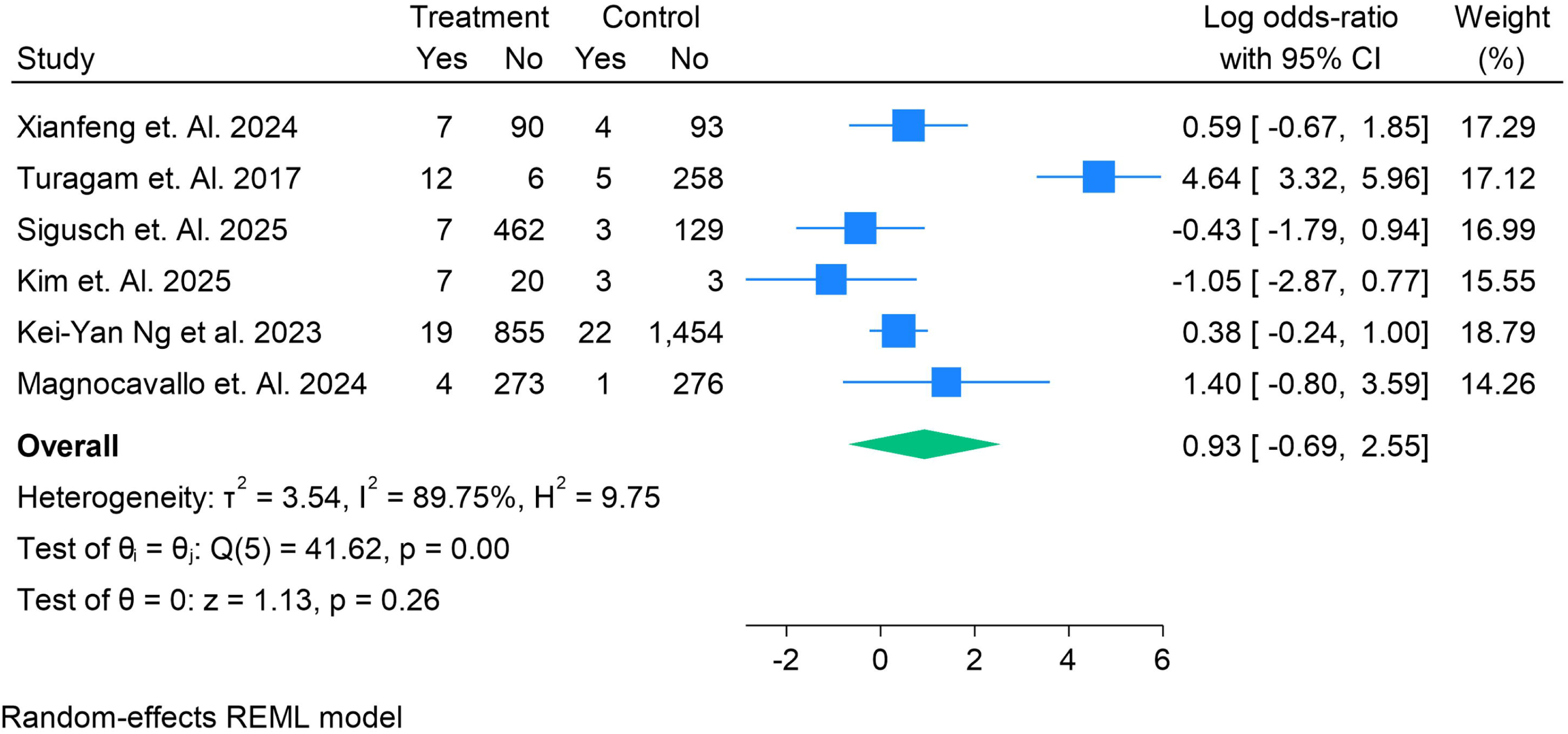
The Odds of the sample having the Pericardia! Effusion in LAAO vs DOACs

#### Vascular Complications

Five studies reported vascular complications in patients undergoing LAAO compared with those treated with DOACs. The pooled analysis using a random-effects REML model showed **no significant difference** between groups (log odds ratio: 0.09; 95% CI: –0.65 to 0.83; p = 0.81). Statistical heterogeneity was low (I² = 34.9%, τ² = 0.24, p = 0.15), suggesting reasonable consistency across studies. Most individual trials, including Paiva et al. (2021), Xianfeng et al. (2024), and Nielsen-Kudsk et al. (2021), demonstrated no significant differences. Kei-Yan Ng et al. (2023) reported a modest increase in vascular complications with LAAO (log OR 1.38; 95% CI: 0.02–2.73), though this effect was not replicated elsewhere. Overall, these findings indicate that **vascular complication rates are similar between LAAO and DOAC therapy**, with no consistent evidence of increased procedural vascular risk in the LAAO group. **Figure 8**. Forest plot showing the odds of vascular complications in patients treated with LAAO versus DOAC therapy. The square sizes reflect study weights, and horizontal lines represent 95% confidence intervals. Pooled estimates were obtained using a random-effects REML model.

**Figure 8.**
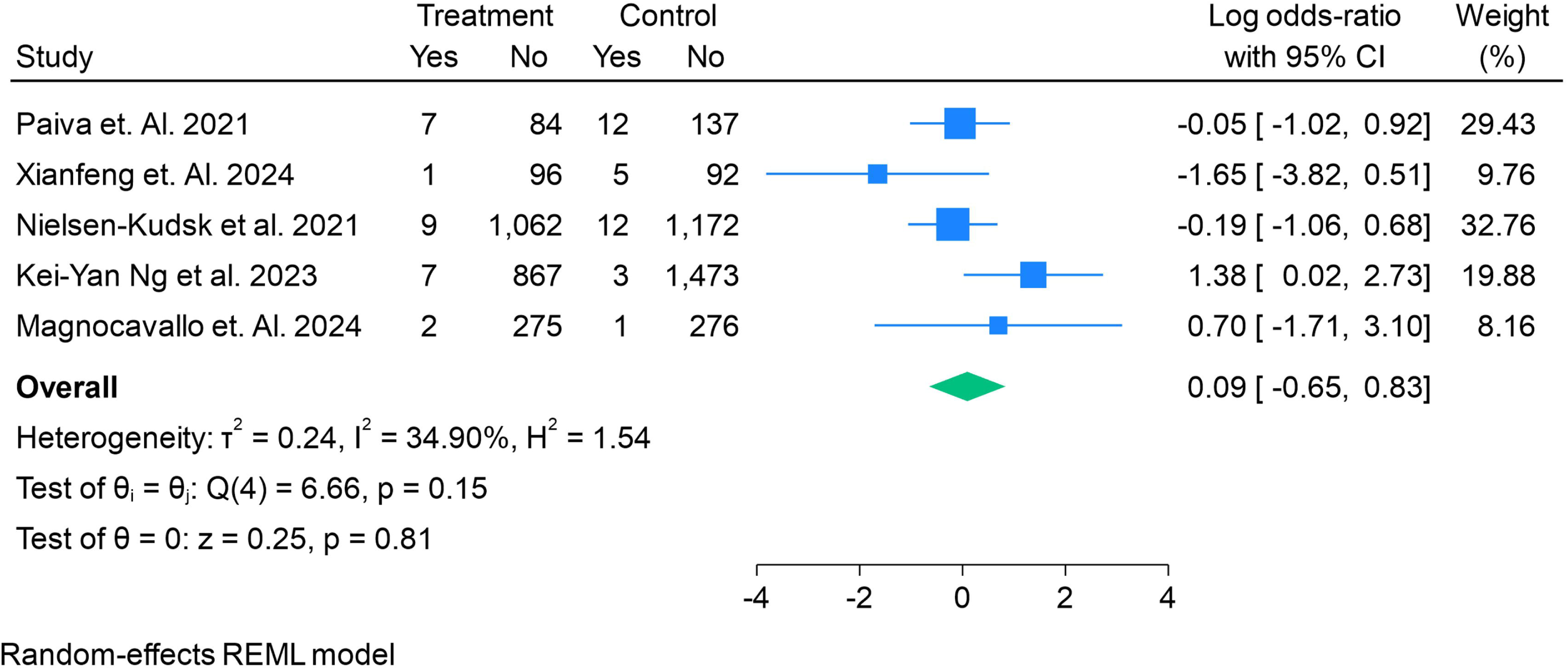
The Odds of the sample having the vascular complication in LAAO vs DOACs

#### Cardiovascular Mortality

Nine studies reported on cardiovascular (CV) mortality in patients treated with LAAO compared with DOAC therapy. The pooled analysis using a random-effects REML model demonstrated **no significant difference** between the two groups (log risk ratio: 0.67; 95% CI: –0.60 to 1.94; p = 0.30). However, **very high heterogeneity** was present (I² = 98.4%, τ² = 3.43, p < 0.001). While several studies, including Nielsen-Kudsk et al. (2021) and Kei-Yan Ng et al. (2023), showed a lower risk of CV mortality with LAAO, others, such as Noseworthy et al. (2022) and Ding et al. (2022), demonstrated markedly higher mortality in the LAAO group. Aarnik et al. (2024) and Magnocavallo et al. (2024) reported no significant differences between groups. These findings suggest that **the effect of LAAO versus DOACs on CV mortality remains uncertain**, with substantial variation likely driven by differences in patient selection, study design, and follow-up duration. **Figure 9**. Forest plot showing the risk ratios of cardiovascular mortality in patients undergoing LAAO compared with DOAC therapy. Square sizes reflect study weight, and horizontal lines represent 95% confidence intervals. Pooled effects were calculated using a random-effects REML model.

**Figure 9.**
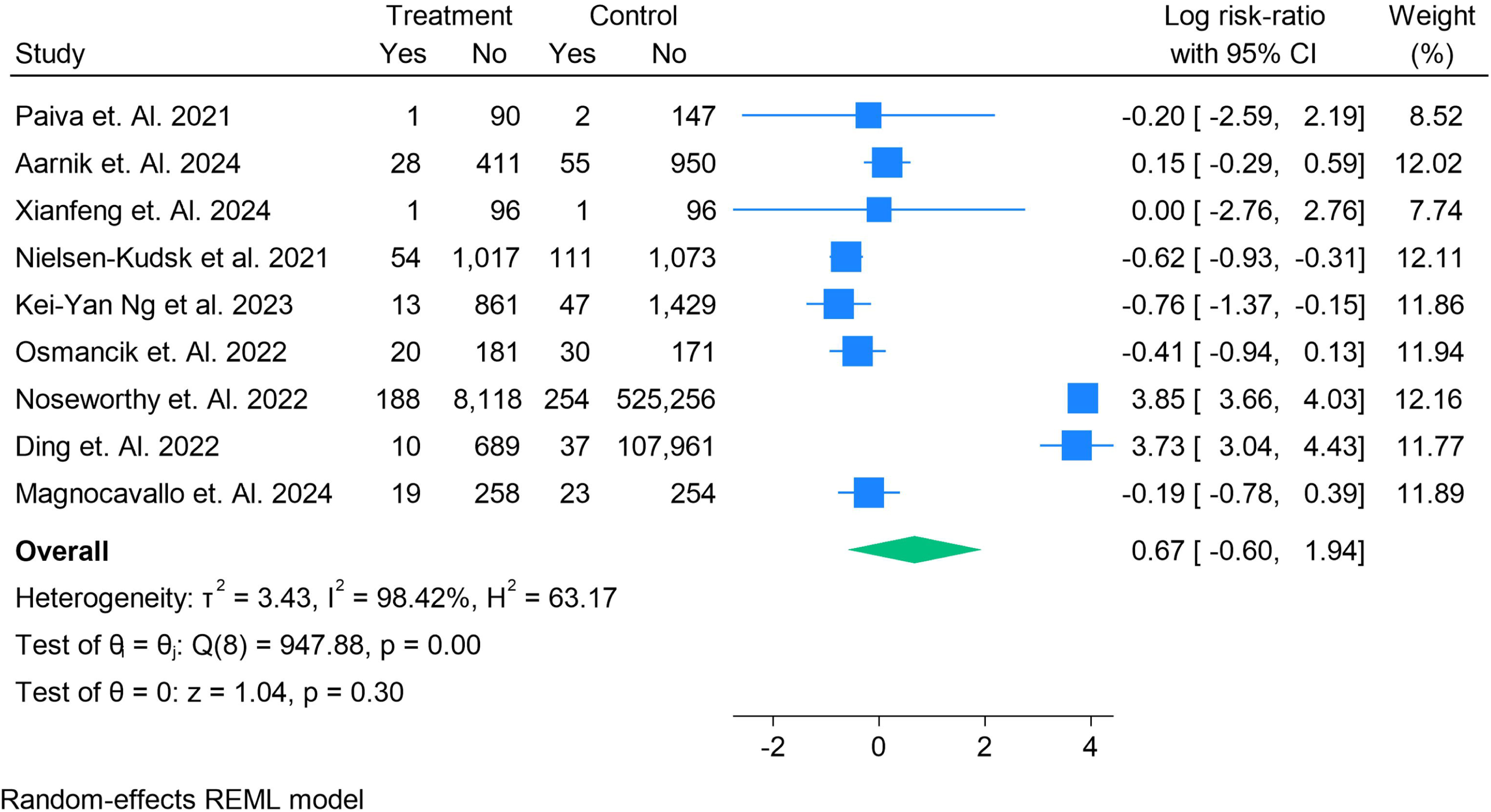
The Risk Ratios of CV Mortality in LAAO vs DOACs

#### All-Cause Mortality

Ten studies reported all-cause mortality among patients undergoing LAAO compared with those receiving DOAC therapy. The pooled analysis using a random-effects REML model found **no significant difference** between groups (log risk ratio: 0.08; 95% CI: –0.77 to 0.92; p = 0.86). There was, however, **very high heterogeneity** (I² = 97.7%, τ² = 1.69, p < 0.001). Nielsen-Kudsk et al. (2021) and Kei-Yan Ng et al. (2023) demonstrated lower all-cause mortality with LAAO, while Ding et al. (2022) reported substantially higher mortality in LAAO patients. Most other studies, including Paiva et al. (2021), Aarnik et al. (2024), and Magnocavallo et al. (2024), found no significant differences between groups. Overall, these results indicate that **the effect of LAAO on all-cause mortality compared with DOAC therapy remains inconclusive**, with study-level differences likely reflecting variations in patient selection, risk profile, and length of follow-up. **Figure 10**. Forest plot of risk ratios for all-cause mortality in patients treated with LAAO versus DOAC therapy. The square sizes represent study weights, and horizontal lines indicate 95% confidence intervals. The pooled effect estimate was calculated using a random-effects REML model.

**Figure 10.**
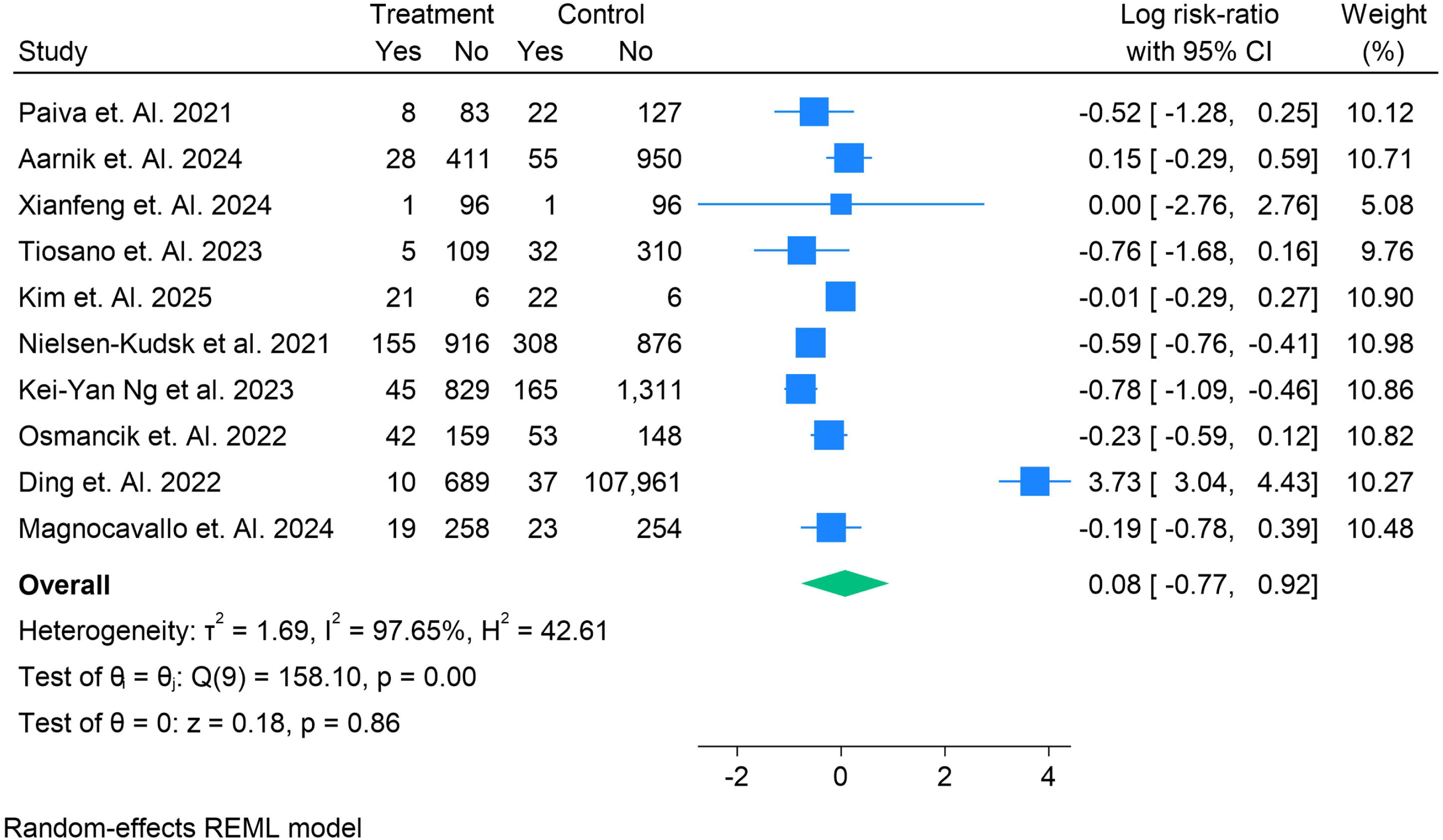
The Risk Ratios of All Cause Mortality in LAAO vs DOACs

#### Hemorrhagic Shock

Five studies assessed the risk of hemorrhagic shock among patients undergoing LAAO compared with those treated with DOACs. The pooled analysis using a random-effects REML model showed no significant difference between groups (log risk ratio: 0.01; 95% CI: –0.31 to 0.32; p = 0.97). Notably, there was no heterogeneity (I² = 0.0%, τ² = 0.00, p = 0.59), indicating highly consistent findings across studies. Individual studies, including Nielsen-Kudsk et al. (2021), Osmancik et al. (2022), and Kei-Yan Ng et al. (2023), demonstrated nearly identical risks between LAAO and DOAC therapy. Tiosano et al. (2023) and Kim et al. (2025) also reported no meaningful differences, with wide confidence intervals reflecting smaller sample sizes. Overall, these results indicate that LAAO does not increase the risk of hemorrhagic shock compared with DOAC therapy, and consistency across studies reinforces the robustness of this finding. Figure 11. Forest plot showing the risk ratios of hemorrhagic shock in patients undergoing LAAO compared with DOAC therapy. The square sizes represent study weights, and horizontal lines denote 95% confidence intervals. Pooled estimates were derived using a random-effects REML model.

**Figure 11.**
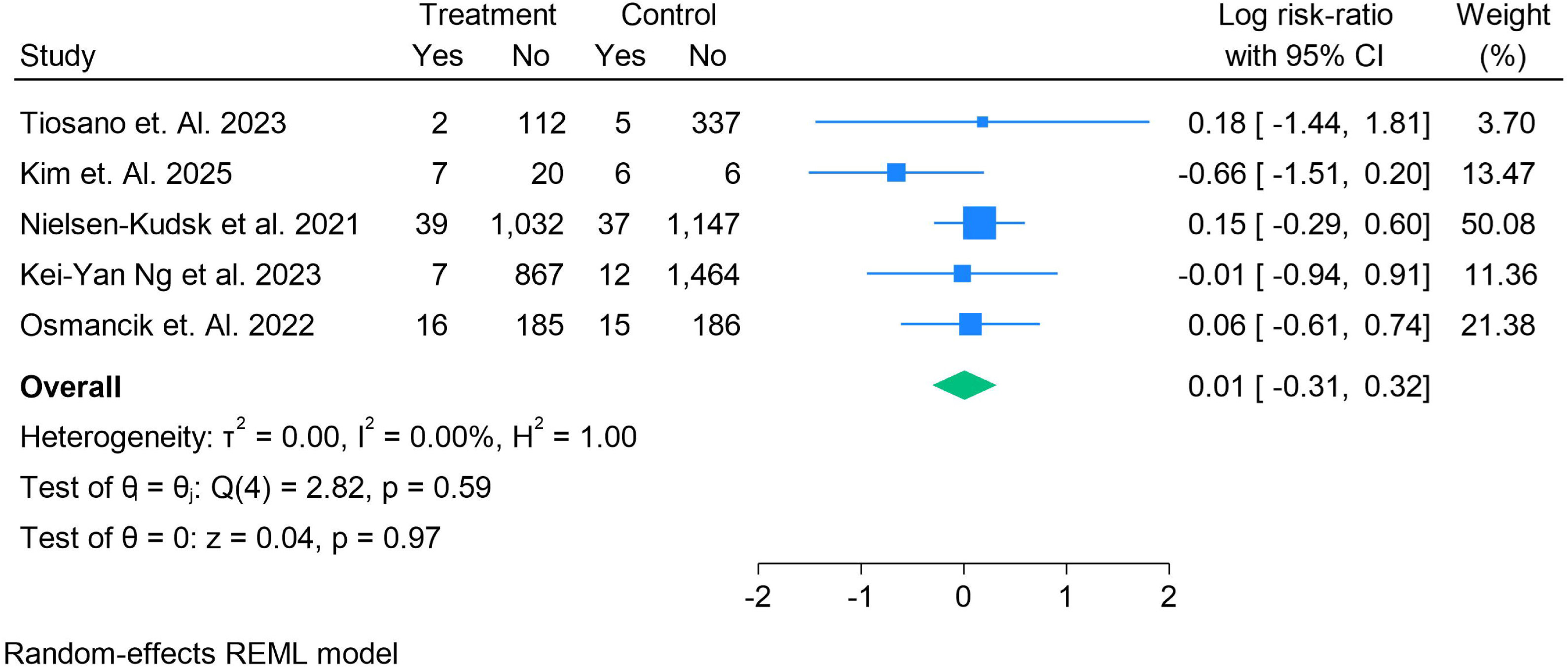
The Risk Ratios of Hemorrhagic Shock in LAAO vs DOACs

#### Thromboembolic Events

Nine studies reported on thromboembolic events in patients undergoing LAAO compared with those receiving DOAC therapy. The pooled analysis using a random-effects REML model demonstrated **no significant difference** between the two strategies (log risk ratio: 0.47; 95% CI: –0.83 to 1.77; p = 0.48). There was **very high heterogeneity** (I² = 98.5%, τ² = 3.64, p < 0.001). Results varied considerably across studies: Ding et al. (2022) and Khalid et al. (2024) reported significantly higher event rates with LAAO, whereas Xianfeng et al. (2024) showed a lower risk. Most other studies, including Nielsen-Kudsk et al. (2021), Paiva et al. (2021), and Osmancik et al. (2022), demonstrated no significant differences. Overall, these findings suggest that **LAAO and DOAC therapy offer broadly comparable protection against thromboembolic events**, but substantial variability across studies highlights the influence of patient selection, anticoagulation strategies post-procedure, and differences in follow-up duration. **Figure 12**. Forest plot showing risk ratios of thromboembolic events in patients treated with LAAO compared with DOAC therapy. Squares represent study weights, and horizontal lines indicate 95% confidence intervals. The pooled effect estimate was generated using a random-effects REML model.

**Figure 12.**
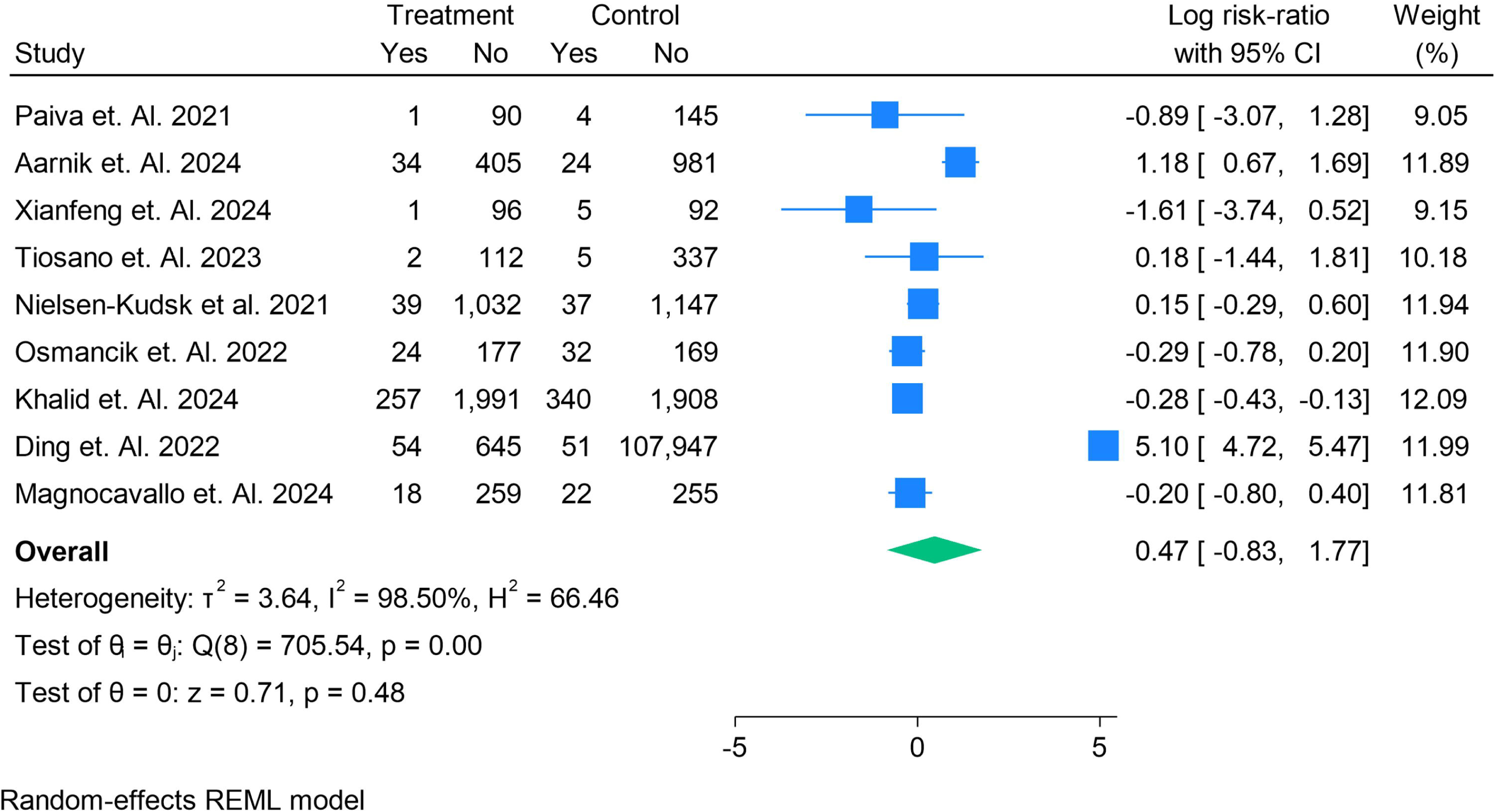
The Risk Ratios ofThromboembolic Events in LAAO vs DOACs

#### Stroke

Ten studies reported stroke outcomes in patients treated with LAAO compared with DOAC therapy. The pooled analysis using a random-effects REML model found **no significant difference** between the two strategies (log risk ratio: 0.97; 95% CI: –0.29 to 2.22; p = 0.13). There was **very high heterogeneity** across studies (I² = 99.4%, τ² = 3.85, p < 0.001). Some studies, such as Ding et al. (2022) and Noseworthy et al. (2022), demonstrated substantially higher stroke risk in the LAAO group, while others, including Nielsen-Kudsk et al. (2021), Kei-Yan Ng et al. (2023), and Magnocavallo et al. (2024), showed no significant differences. Aarnik et al. (2024) suggested a modest increase in risk, whereas Khalid et al. (2024) reported a protective effect of LAAO. Overall, these findings indicate that **stroke rates appear broadly comparable between LAAO and DOAC therapy**, but substantial heterogeneity highlights differences in study design, baseline risk, and follow-up duration. **Figure 13**. Forest plot showing risk ratios of stroke in patients treated with LAAO compared with DOAC therapy. The size of the squares represents study weight, and horizontal lines indicate 95% confidence intervals. Pooled estimates were generated using a random-effects REML model.

**Figure 13.**
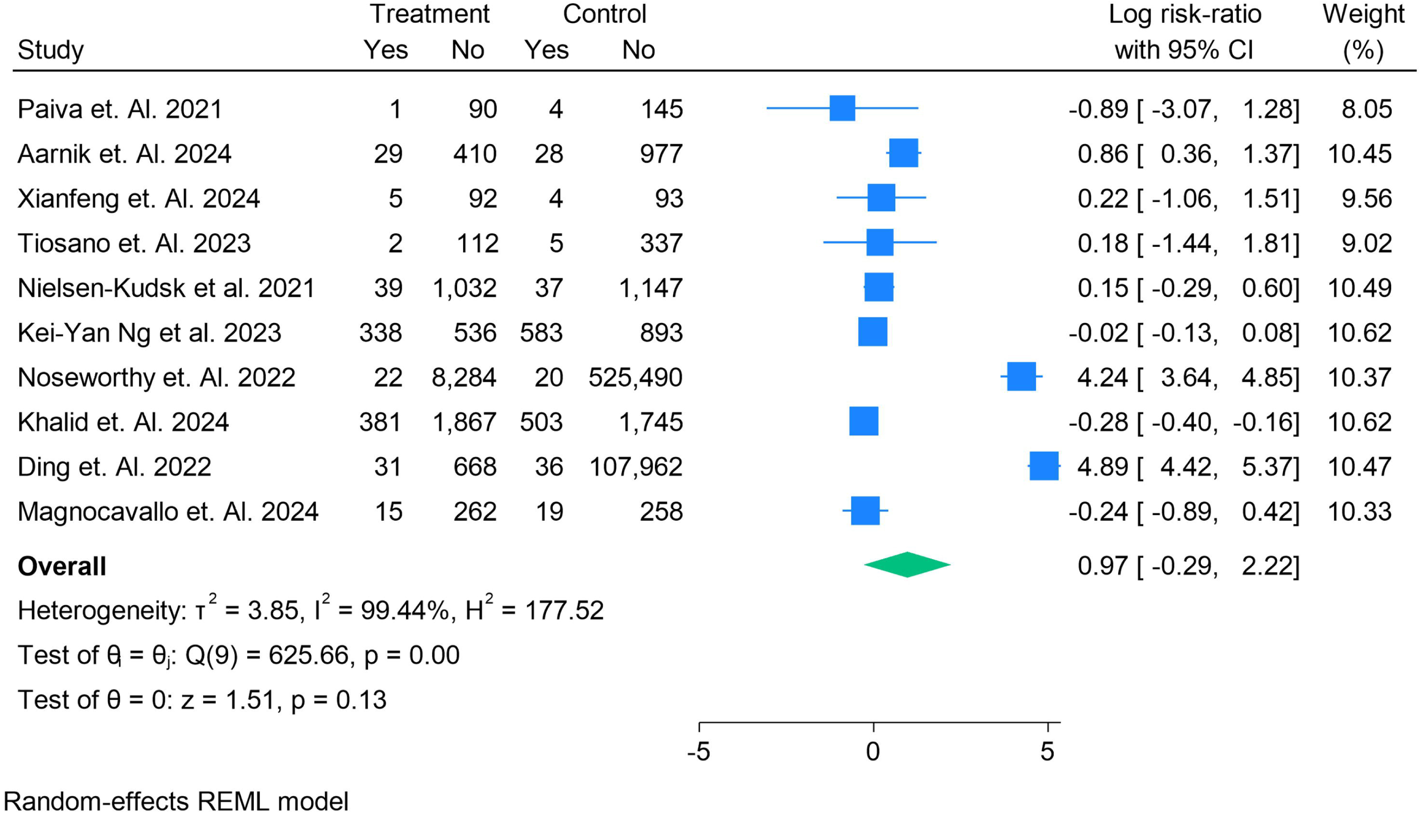
The Risk Ratios of Stroke in LAAO vs DOACs

#### Bleeding Events

Eleven studies reported on bleeding outcomes in patients undergoing LAAO compared with those treated with DOACs. The pooled analysis using a random-effects REML model demonstrated **no significant difference** between the two groups (log risk ratio: 0.58; 95% CI: –0.71 to 1.87; p = 0.38). However, there was **very high heterogeneity** (I² = 99.6%, τ² = 4.49, p < 0.001). Results varied substantially across studies: Noseworthy et al. (2022) and Ding et al. (2022) reported markedly higher bleeding rates with LAAO, whereas Nielsen-Kudsk et al. (2021) and Khalid et al. (2024) suggested lower risks. Other studies, including Paiva et al. (2021), Aarnik et al. (2024), and Magnocavallo et al. (2024), reported mixed or neutral findings. Taken together, these findings indicate that **LAAO and DOACs yield broadly comparable bleeding outcomes**, but wide heterogeneity underscores differences in patient characteristics, antithrombotic regimens, and bleeding definitions across studies. **Figure 14**. Forest plot showing risk ratios of bleeding in patients treated with LAAO compared with DOAC therapy. Squares represent study weights, and horizontal lines indicate 95% confidence intervals. Pooled estimates were calculated using a random-effects REML model.

**Figure 14.**
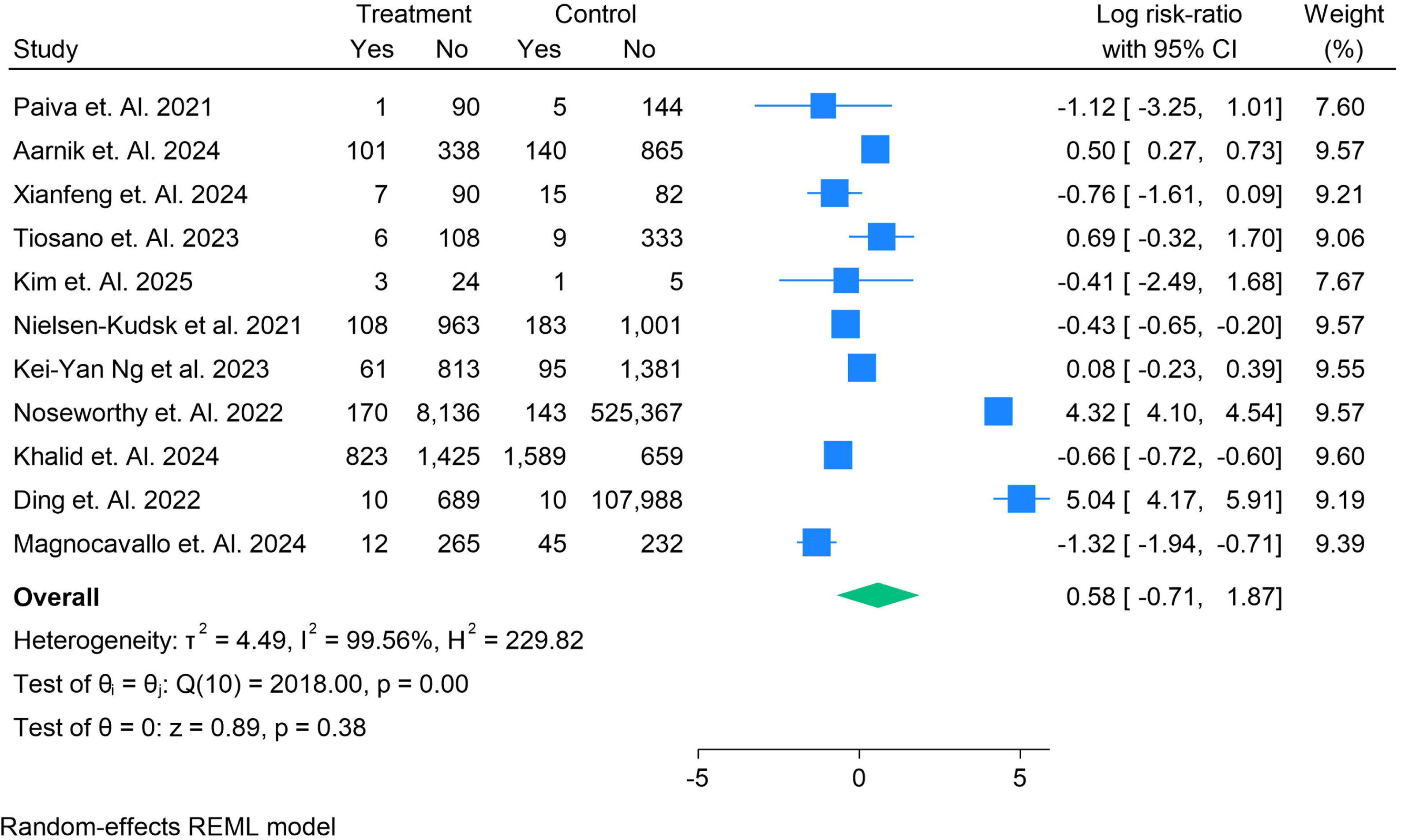
The Risk Ratios of Bleeding in LAAO vs DOACs

## Discussion

Our meta-analysis, synthesizing data from over 680,000 patients across 14 studies, provides an updated comparison of left atrial appendage occlusion (LAAO) and direct oral anticoagulants (DOACs) in patients with atrial fibrillation. This is one of the most comprehensive assessments to date, combining randomized evidence with large-scale observational data, thereby offering insights into real-world effectiveness and safety of these two approaches to stroke prevention. The principal finding of this analysis is that LAAO and DOAC therapy demonstrate broadly comparable outcomes with respect to all-cause mortality, cardiovascular mortality, thromboembolic events, and major bleeding, although individual study results varied considerably and heterogeneity was substantial in many endpoints.

With regard to mortality, pooled results showed no significant difference in either all-cause or cardiovascular death between the two strategies. Some studies, such as Nielsen-Kudsk et al. and Ng et al., suggested improved survival with LAAO, whereas registry data from Noseworthy et al. and Ding et al. indicated higher event rates in the LAAO arm. This inconsistency likely reflects confounding by indication, where LAAO is often reserved for patients at highest baseline risk, particularly those with bleeding or renal dysfunction that preclude long-term anticoagulation. Such patients may inherently carry worse outcomes, regardless of the intervention. Therefore, mortality comparisons should be interpreted with caution, and only adequately powered randomized trials can definitively address this issue.

Analysis of thromboembolic events, including ischemic stroke and systemic embolism, revealed no consistent difference between LAAO and DOACs. Several studies showed equivalence, while some suggested a transiently higher ischemic risk following LAAO, particularly in the peri-procedural period. This may be attributable to incomplete device endothelialization, peridevice leaks, or early discontinuation of adjunctive antithrombotic therapy. Over longer follow-up, however, outcomes appear comparable, supporting the concept that once procedural risk is overcome, LAAO can provide durable protection similar to DOACs. These results are consistent with prior randomized trials such as PROTECT-AF and PREVAIL, which established non-inferiority of LAAO against warfarin.

Bleeding outcomes were complex and heterogeneous. Overall, there was no significant difference in major bleeding between groups, though heterogeneity exceeded 90%, reflecting variability in study design, follow-up, and definitions of bleeding endpoints. Procedural and device-related complications, such as pericardial effusion and vascular injury, were predictably more frequent in LAAO recipients, emphasizing the importance of procedural expertise and device iteration. Conversely, some long-term data suggest that LAAO reduces the incidence of late bleeding, particularly intracranial and gastrointestinal hemorrhage, by eliminating the need for chronic anticoagulation. Observational studies such as those by Nielsen-Kudsk and Khalid demonstrated lower bleeding rates with LAAO in selected populations, although these findings were not universally reproduced. Importantly, pooled analyses of ISTH-defined bleeding, hemorrhagic shock, and BARC outcomes indicated no clear excess risk in either group.

Safety findings in this review highlight that patient selection is critical. For individuals with absolute contraindications to anticoagulation, recurrent bleeding, or advanced chronic kidney disease, LAAO offers a reasonable and guideline-supported alternative. For the majority of patients, however, DOACs remain the first-line therapy given their robust evidence base, predictable pharmacokinetics, and proven mortality benefit. Procedural risks of LAAO, though diminishing with newer-generation devices such as WATCHMAN FLX and Amulet, remain non-trivial and must be balanced against the risks of long-term anticoagulation. Importantly, as techniques mature and operator experience expands, peri-procedural complication rates are expected to continue declining, potentially shifting the balance in favor of LAAO in carefully selected patients.

When placed in the context of previous systematic reviews, our findings are largely consistent with recent meta-analyses that demonstrated non-inferiority of LAAO compared with oral anticoagulation [34]. Most prior reviews, however, compared LAAO with vitamin K antagonists rather than DOACs. Our study extends the literature by focusing specifically on DOACs, the current standard of care, and by including the most contemporary data up to September 2025. The absence of significant differences in efficacy or safety underscores that LAAO should not be viewed as a blanket replacement for DOACs but rather as a complementary strategy for specific high-risk subsets.

Several limitations must be acknowledged. The majority of included studies were observational, and despite statistical adjustment, residual confounding cannot be excluded [35]. Heterogeneity across studies was high, reflecting differences in patient selection, endpoint definitions, follow-up duration, and device types. Early-generation devices were associated with higher complication rates, making it difficult to generalize findings to current practice with newer technology. Furthermore, the lack of individual patient-level data prevented detailed subgroup analyses to identify which patients benefit most from LAAO. Finally, publication bias may exist, as studies with neutral or negative outcomes may be underrepresented.

## Conclusion

In conclusion, this meta-analysis demonstrates that LAAO and DOAC therapy yield comparable outcomes for stroke prevention in atrial fibrillation, with no consistent differences in mortality or major bleeding. LAAO remains a valuable option for patients unable to tolerate long-term anticoagulation, particularly those at high bleeding risk, while DOACs continue to represent the standard of care for the broader AF population. Future large-scale randomized trials directly comparing LAAO with DOACs are warranted to refine patient selection, confirm long-term efficacy, and define the role of this strategy in contemporary AF management.

## Conflict of Interest

*The authors certify that there is no conflict of interest with any financial organization regarding the material discussed in the manuscript*.

## Funding

*The authors report no involvement in the research by the sponsor that could have influenced the outcome of this work*.

## Authors’ contributions

*Manish Juneja conceptualized the study, provided overall supervision, and critically revised the manuscript. Deep Patel contributed to literature search, data extraction, and drafting of the initial manuscript. Manasi Garde assisted in data analysis, interpretation of results, and manuscript editing. Aadhiti Kandula, parag n patel contributed to data acquisition, tabulation, and reference management. Maria Elizabeth John performed quality checks of extracted data and supported in manuscript formatting. Goranti Shreya Reddy contributed to statistical analysis and figure preparation. Vivek Shekar assisted in drafting the discussion section and proofreading. Rakhshanda Khan provided methodological guidance, contributed to data interpretation, and supervised the overall writing process. Harshawardhan Dhanraj Ramteke coordinated project administration, contributed to study design, and manuscript revisions. Syeda Hafsa Noor-Ain assisted in reviewing the final draft and ensured consistency of reporting*.

*All authors contributed significantly to the work, approved the final version of the manuscript, and agree to be accountable for its content*.

## Data Availability

supplementary file

## Notes

### Competing Interest Statement

The authors have declared no competing interest.

### Clinical Protocols

https://www.crd.york.ac.uk/PROSPERO/view/CRD420251152538

### Funding Statement

none

