## supplementary file for "Comparative Efficacy and Safety of Left Atrial Appendage Occlusion Versus Direct Oral Anticoagulants in Atrial Fibrillation: A Systematic Review and Meta-Analysis"

Supplementary Files

Search String

(("left atrial appendage occlusion"[Title/Abstract] OR "LAA occlusion"[Title/Abstract] OR "LAAO"[Title/Abstract] OR "watchman device"[Title/Abstract] OR "Amplatzer Amulet"[Title/Abstract] OR "LAAC"[Title/Abstract])

AND

("direct oral anticoagulant*"[Title/Abstract] OR "DOAC*"[Title/Abstract] OR "novel oral anticoagulant*"[Title/Abstract] OR "NOAC*"[Title/Abstract] OR "dabigatran"[Title/Abstract] OR "rivaroxaban"[Title/Abstract] OR "apixaban"[Title/Abstract] OR "edoxaban"[Title/Abstract])

AND

("atrial fibrillation"[Title/Abstract] OR "AF"[Title/Abstract]))

Table S1. Findings of Studies

| **Author Year** | **Country** | **Type of Study** | **Follow Up Month** | **Total Sample** | **Male** | **Female** | **LAAO Number** | **AF Number** | **HTN in LAAO** | **HTN in DOAC** | **CKD in LAAO** | **CKD in DOAC** | **Diabetes** | **Diabetes** | **Heart Failure** | **Heart Failure** | **chads2vasc** | **chads2vasc** | **HASBLED Score** | **HASBLED Score** | **LAAO Device** | **Grade** | **Main Findings** |
| --- | --- | --- | --- | --- | --- | --- | --- | --- | --- | --- | --- | --- | --- | --- | --- | --- | --- | --- | --- | --- | --- | --- | --- |
| Paiva et. Al. 2021 | Portugal | Prospective, single-center, non-randomized cohort stud | 13 | 240 | 128 | 112 | 91 | 149 | 79 | 123 | 25 | 48 | 32 | 36 | 42 | 34 | 4.3±1.4 | 5.3±1.3 | 3.0±0.9 | 4.0±0.7 | Watchman | Moderate | LAAO was not inferior to NOAC for preventing all-cause mortality, stroke, and major bleeding in high-risk atrial fibrillation patients |
| Aarnik et. Al. 2024 | Multicenter | Multicenter Retrospective | 28 | 1444 | 265 | 601 | 439 | 1005 | 336 | 364 | 40 | 56 | 108 | 117 | 104 | 122 | 5.0 ± 1.6 | 5.2 ± 1.6 | 2.8 ± 1.3 | 2.4 ± 1.2 | Watchman | High | *LAAO in patients with anticoagulation failure showed similar stroke prevention efficacy compared with LAAO in anticoagulation-contraindicated patients, with fewer bleeding events but higher thromboembolic risk* |
| Xianfeng et. Al. 2024 | Multicenter | Prospective, multicenter, randomized clinical trial | 30 | 202 | 110 | 84 | 97 | 97 | 58 | 58 |  |  | 28 | 25 | 17 | 20 | 4 ± 1.6 | 4 ± 1.6 | 2 ± 1.8 | 2 ± 1.8 | Watchman | Moderate | Occlusion-first strategy reduced thromboembolic events, device thrombus, and improved long-term arrhythmia-free survival compared with ablation-first |
| Turagam et. Al. 2017 | USA | Multicenter, **retrospective observational study** | 14 | 263 | 95 | 168 | 18 | 263 | 145 |  | 12 |  | 42 |  | 21 |  | **3.2 ± 1.6** |  | 3.8 ± 0.8 |  | Watchman | Moderate | In warfarin-ineligible AF patients re-challenged with DOACs, repeat major bleeding was frequent (63%), while LAAO devices provided safe outcomes in high-risk patients requiring major interventions |
| Tiosano et. Al. 2023 | Israel | Indirect, retrospective comparison of two prospective registries | 12 | 456 | 272 | 184 | 114 | 342 | 98 | 279 | 62 | 217 | 50 | 137 | 44 | 124 | 4.17 ± 1.29 | 3.96 ± 1.57 | 4.14 ± 1.04 | 3.95 ± 1.35 | Amplatzer Amulet | Moderate | LAAO was associated with significantly lower 1-year all-cause mortality compared with NOACs, especially in patients with impaired renal functio |
| Sigusch et. Al. 2025 | Germany | Single-center, retrospective **observational study** | 25 | 599 | 369 | 230 | 469 | 132 |  |  |  |  |  |  |  |  | 4.5 ± 1.4 |  | 3.2 ± 1.0 |  | Watchman | Moderate | *LAAO increased hemoglobin concentration in post-bleeding patients, while high-bleeding-risk patients showed a decline, with overall acceptable safety outcomes* |
| Kim et. Al. 2025 | USA | Retrospective single-center **observational case series** | 6 | 33 | 23 | 10 | 27 | 6 | 22 | 6 | 1 | 1 | 8 | 4 | 17 | 5 | 4.6 ± 1.4 | **6.5 ± 0.5** | 4.3 ± 0.8 | 4.7 ± 0.5 | Watchman | Moderate | *Patients with lower CHA₂DS₂-VASc scores were more likely to receive minimal therapy post-LAAO, with no significant difference in bleeding or thromboembolic events compared to standard therapy.* |
| Nielsen-Kudsk et al. 2021 | Danish | Observational cohort study | 24 | 1071 | 1184 | 2255 | 1071 | 1184 | 896 | 1023 | 149 | 169 | 333 | 424 |  |  | 4.2 ± 1.6 | 4.3 ± 1.7 | 3.3 ± 1.0 | 3.4 ± 1.2 | Amplatzer Amulet | Moderate | *LAAO provided similar stroke prevention but significantly reduced major bleeding, cardiovascular and all-cause mortality compared with DOACs in high-risk AF patients* |
| Kei-Yan Ng et al. 2023 | China | Retrospective cohort | 35 | 2350 | 1349 | 1001 | 874 | 1476 | 586 | 1004 | 48 | 70 | 263 | 447 | 235 | 449 | 4.4 ± 1.7 | 4.5 ± 1.7 | 4.6 ± 1.23 | 4.2 ± 1.23 | Watchman | Moderate | LAAO showed **similar composite outcomes** to DOAC switch but significantly **reduced all-cause and cardiovascular mortality** and **lower bleeding after 6 months** |
| Osmancik et. Al. 2022 | Czech Republic | Randomized, prospective, open-label, multicenter, noninferiority trial | 42 | 402 | 264 | 138 | 201 | 201 | 186 | 186 |  |  | 73 | 90 | 88 | 90 | 4.7 ± 1.5 | 4.7 ± 1.5 | 3.1 ± 0.9 | 3.0 ± 0.9 | Watchman | Moderate | *LAAC was noninferior to DOACs for major cardiovascular, neurological, or bleeding events, with significantly less long-term nonprocedural bleeding* |
| Noseworthy et. Al. 2022 | Atrial Fibrillation | Retrospective observational | 18 | 562852 | 280097 | 282753 | 8306 | 525510 | 2912 | 120954 | 2912 | 120954 | 4904 | 258544 | 5220 | 237091 | 5.9 ± 1.6 | 5.1 ± 1.8 |  |  | Watchman | Moderate | LAAO had similar composite outcomes as NOACs, with lower all-cause mortality but higher major bleeding risk. |
| Khalid et. Al. 2024 | USA | Retrospective, multicenter, **1:1 propensity-matched cohort study** | 60 | 4496 | 2340 | 2156 | 2248 | 2248 | 2187 | 2199 | 1000 | 1000 | 1228 | 1228 | 41 | 48 | 4.81 ± 1.25 | 4.81 ± 1.25 |  |  | Watchman | Moderate | *The Watchman device reduced long-term bleeding, ischemic stroke, and TIA compared to DOACs, with similar hemorrhagic stroke rates over 5 years.* |
| Ding et. Al. 2022 | Spain | Observational, retrospective cohort study | 24 | 108697 | 240 | 49718 | 699 | 107998 | 496 | 63165 |  |  | 246 | 23695 | 257 | 20576 |  |  |  |  | Watchman | Moderate | *LAAO was associated with significantly lower all-cause mortality compared to DOACs, with no significant difference in thromboembolic or bleeding outcomes* |
| Magnocavallo et. Al. 2024 | USA | Prospective, multicenter, observational cohort with **propensity-score matching**. | 25 | 554 | 295 | 408 | 277 | 277 | 261 | 266 | 7 | 6 | 107 | 114 | 130 | 149 | 5.8 ± 0.9 | 5.8 ± 0.9 | 3.0 ± 0.8 | 3.0 ± 0.9 | Watchman | Moderate | **LAAO in very high-risk AF patients provided similar stroke prevention but significantly reduced clinically relevant bleeding compared to DOACs, with benefits emerging after ~18 months.** |

Robins

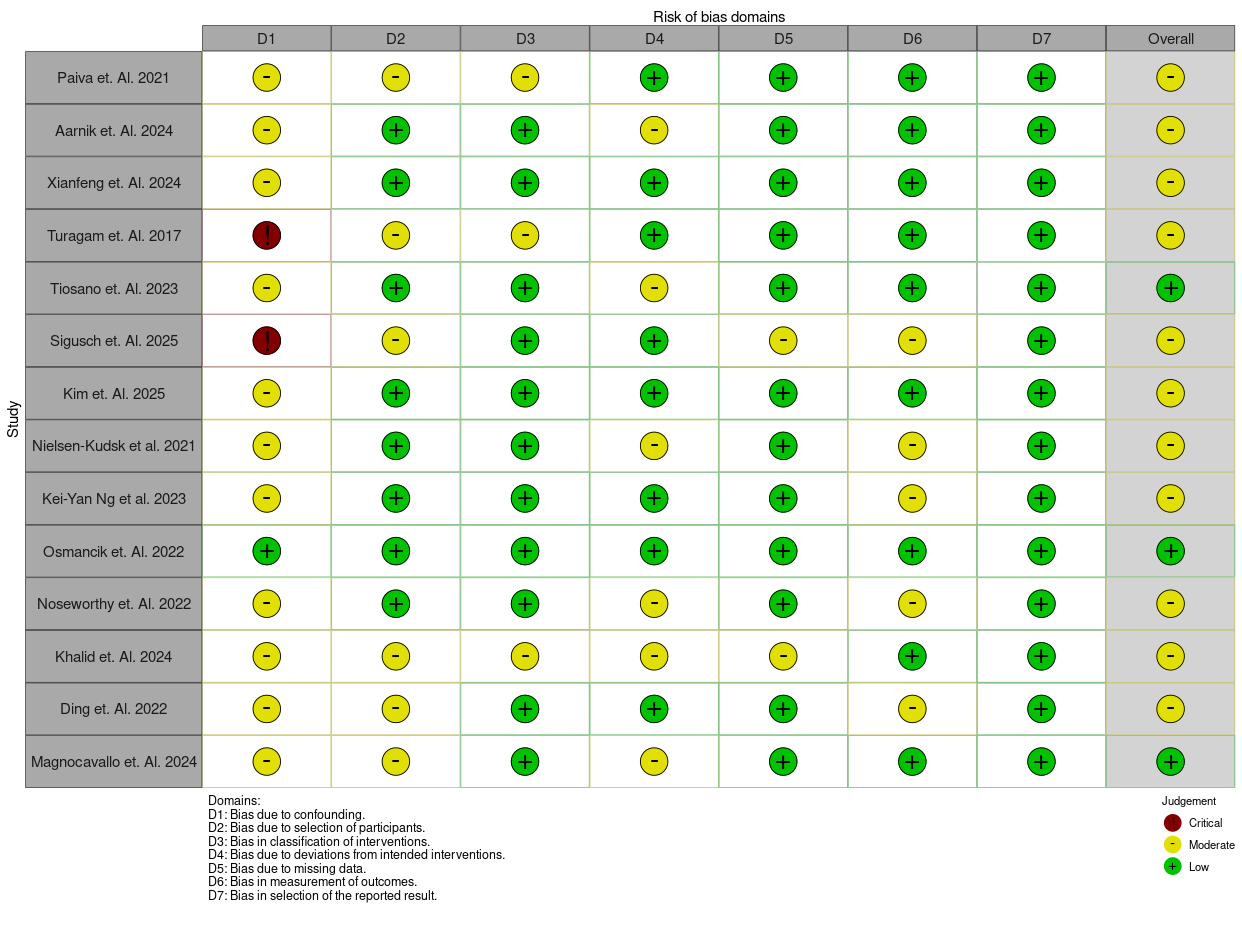
